# Mild Adverse Events of Sputnik V Vaccine Extracted from Russian Language Telegram Posts via BERT Deep Learning Model

**DOI:** 10.1101/2021.06.14.21258875

**Authors:** Andrzej Jarynowski, Alexander Semenov, Mikołaj Kamiński, Vitaly Belik

**Affiliations:** System Modeling Group, Institute of Veterinary Epidemiology and Biostatistics, Freie Universität Berlin, Berlin, Germany; Herbert Wertheim College of Engineering, University of Florida, FL, USA; Center for Econometrics and Business Analytics. St. Petersburg State University, Russia; Individual Medical Practice, Bogdanowo, 64-600 Oborniki, Poland

**Keywords:** adverse events, Sputnik V, Gam-COVID-Vac, social media, Telegram, COVID19, Sars-CoV-2, deep learning

## Abstract

**Background:** There is a limited amount of data on the COVID-19 vector vaccine Gam-COVID-Vac (Sputnik V) safety profile. Previous infodemiology studies showed that social media discourse could be analyzed to assess the most concerning adverse events (AE) caused by drugs.

**Objective:** We aimed to investigate mild AEs of Sputnik V based on a participatory trial conducted on Telegram in the Russian language. We compared AEs extracted from Telegram with other limited databases on Sputnik V and other COVID-19 vaccines. We explored symptom co-occurrence patterns and determined how counts of administered doses, age, gender, and sequence of shots could confound the reporting of AEs.

**Materials and Methods:** We collected a unique dataset consisting of 11,515 self-reported Sputnik V vaccine AEs posted on the Telegram group, and we utilized natural language processing methods to extract AEs. Specifically, we performed multi-label classifications using the deep neural language model BERT “DeepPavlov”, which we pre-trained on a Russian language corpus and applied to the Telegram messages. The resulting AUC score was 0.991. We chose symptom classes that represented the following AEs: fever, pain, chills, fatigue, nausea/vomiting, headache, insomnia, lymph node enlargement, erythema, pruritus, swelling, and diarrhea.

**Results:** The results of the retrospective analysis showed that females reported more AEs than males (1.2-fold, P<.001). In addition, there were more AEs from the first dose than from the second dose (1.13-fold, P<.001), and the number of AEs decreased with age (*β* = .05 per year, P<.001). The results also showed that Sputnik V AEs were more similar to other vector vaccines (132 units) compared with mRNA ones (241 units) according to the average Euclidean distance between the vectors of AE frequencies. Elderly Telegram users reported significantly more (5.6-fold on average) systemic AEs than their peers, according to the results of the phase III clinical trials published in *The Lancet*. However, the AEs reported in Telegram posts were consistent (Pearson correlation r=.94, P=.02) with those reported in the Argentinian post-marketing AE registry.

**Conclusion:** After receiving the Sputnik V vaccination, Telegram users complained about pain (47%), fever (47%), fatigue (34%), and headache (25%). The results showed that the AE profile of Sputnik V was comparable with other COVID-19 vaccines. Examining the sentinel properties of participatory trials (which is subject to self-reporting biases) could still provide meaningful information about pharmaceutics, especially if only a limited amount of information on AEs is provided by producers.

## Introduction

The current COVID-19 pandemic is one of the most critical global health problems. The main strategies for its mitigation involve both non-pharmaceutical interventions (e.g., testing and contract tracing) and up-to-date anti-COVID-19 treatments. However, the most promising remedy has been vaccines that have effectively prevented severe COVID-19 outcomes. In addition to novel mRNA vaccines, vector vaccines have been developed. One of the first was Gam-COVID-Vac (Sputnik V), which is a viral two-dose vector vaccine based on two human adenoviruses. Each dose contains a different vector: rAd26 and rAd5. This vaccine was developed by the Gamaleya Research Institute of Epidemiology and Microbiology. Sputnik V contains a gene that encodes SARS-CoV-2’s spike (S) protein [1]. To date, two reports of clinical trials have been published. In the first study, of I/II phases involved a total of 76 participants, who were included in the safety analysis [2]. The report on phase III included detailed descriptions of serious and rare adverse events (AE) as well as mild AEs described in individuals [3] older than 60 years. The overall frequency of AEs was mentioned without complete characteristics of the safety profile, such as the co-occurrence of AEs. Mild AEs are common among all vaccines. Extensive fact sheets on AEs, as well as possible adverse reactions, were provided for vaccines trialed under US (FDA - Food and Drug Administration), UK (MHRA - Medicines and Healthcare products Regulatory Agency), or EU (EMA - European Medicines Agency) regulator bodies, which was not the case with Sputnik V. Until 20 March 2021, of the almost 30 million doses manufactured, approximately eight million doses [4] of COVID-19 immunization have been administered in a single dose in Russia [5]. Moreover, the Russian Federation has signed contracts with dozens of countries to deliver 1.4 billion doses at less than 7 EUR per dose for international buyers [6]. Therefore, there is an emerging need to update the information on Sputnik V’s safety profile using post-marketing surveillance. Because a registry of AEs after vaccination with Sputnik V is difficult to access, social media discourse may be an alternate source of information on AEs.

An increasing number of studies have analyzed English-language social media in the context of vaccinations [7] or vaccine-prevented infectious disease [8]. However, only a few similar studies on Russian social media users have been published [9,10]. Accounts of adverse reactions to drugs have been widely extracted from social media [11] in the context of mining consumer reviews on the Internet [12]. To date, most of these studies processed data collected from Twitter [13–22]. Although social media platforms such as Twitter and Facebook are used in Russia, the most popular platforms are VK.com, which has about 97 million active monthly users [23], and Telegram Messenger, which is ranked second in the Russian Appstore, having 500 million active users overall. Developed in Russia, these platforms are much more popular than alternatives such as Facebook or Twitter [23, 24].

Most previous studies on social media vaccine discourse have focused on the personal beliefs of users. For example, Wang et al. developed a framework to detect vaccine AEs mentioned by Twitter users [25]. However, to date, no study has analyzed social media discourse on non-severe AEs in response to COVID-19 vaccines. In the present study, we collected social media (a Telegram group in the Russian language) data to bridge the gap in information on the most prevalent AEs involving Sputnik V. We focused on the most common AEs and established which were the most prevalent, their co-occurrence, and their associations with users’ characteristics [26]. Finally, we compared the AE profile of Sputnik V with those of other approved COVID-19 vaccines.

## Materials

The dataset analyzed in our study was collected retrospectively from the Telegram group^1^, “Sputnik_results”. The data contained no personal information, and the analysis was performed according to the Terms of Service of the platform [27]. Our analysis was completely anonymous and performed in aggregated form. No possible harm to Telegram users was identified. Therefore, the study did not require ethical committee approval.

### Data Description

Originally, Telegram was aiming to provide secure communication, but later functionality was expanded; it added support of public channels, groups, video calls, and many other features. Telegram groups may be public or private. If a group is public, it may be accessed via the Telegram search engine, and every user may read all its content. A main priority claimed by Telegram is security; users’ data are not disclosed, and only the user’s screen name and picture are shown to the public. The largest Telegram channels have millions of subscribers.

The description of the “Sputnik_results” public group states that its main aim is to collect information on AEs regarding the Sputnik V vaccine. Telegram users may post a description of their symptoms. Moderators of the group oversee the messages and verify that they contain only descriptions of AEs; otherwise, the message is deleted.

In the present study, we collected all messages from the “Sputnik_Results” group using Python Telegram Client telethon [28]. We saved only text messages that were posted in the group; users’ personal details were not extracted. In total, we collected 18,833 messages. After filtering messages that contained only pictures, 11,515 messages remained. The first message was sent on 09 December 2020, and the most recent message was sent on 17 April 2021. The dataset contained 25,660 unique lowercase words.

### Adverse Event Classification

The gold standard used to identify adverse reactions is the MedDRA System Organ Class (SOC), which is applied in the EU (EudraVigilance^2^), the US (VAERS^3^), and the UK (MHRA Yellow Card scheme^4^). However, the system uses a specialized medical vocabulary. In our study, because users of social media communicated in colloquial language [11], we chose a simplified FDA classification system [29–31] that was subdivided into two groups: local reactions (i.e., redness, swelling, and pain at the injection site) and systemic reactions (i.e., fever, fatigue, headache, chills, nausea/vomiting, diarrhea, new or worsening muscle pain, and new or worsening joint pain). Moreover, muscle pain, joint pain, and pain at the injection site were categorized as a single class. However, we added the classes of pruritus, enlarged lymph nodes, and insomnia, which are common adverse reactions to anti-COVID-19 vaccines. The final list of 12 classes of symptoms of mild AEs, which were based on subjective experiences of a potential health issue, is provided in Table 2.

## Methods

### Labeling

We utilized the LabelStudio data labeling tool [32] to label the dataset. We randomly sampled 1,000 messages in the dataset, which were labeled by three raters who were native Russian speakers. The raters labeled each occurrence of an AE in the messages, thus making the dataset suitable for named entity recognition (NER) tasks. Because of such labeling and the existence of different descriptions of the same AEs in multiple sentences, we augmented the dataset by splitting each message into sentences. The resulting dataset contained 4,579 entities.

### Model architecture

Each message in our dataset could have included multiple AEs. We therefore adopted a multi-label text classification scheme. A formal definition of multi-label classification is as follows. Consider a dataset, 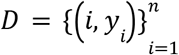 where *x*_*i*_∈*X* is the *i*-th observed variable for the dataset of cardinality *n, y*_*i*_∈*Y* is the corresponding set of labels for the *i*-th element. Our goal was to learn a mapping *ŷ*_*j*_ = *f*(*x*_*i*_, θ), where *ŷ*_*j*_, is the set of predicted classes, and θ is a vector of parameters. To find the vector of optimal parameters θ, we needed to minimize the loss function *L*(*y, ŷ*) between the actual and predicted classes. Multiple machine learning methods may be applied to support multi-label classification. In the case of artificial neural networks (ANN), the activation function of the last layer of the ANN is set to be a sigmoid, and binary cross-entropy loss is used.

Because of the recent success of ANNs, specifically transformers, in text analysis tasks, we adopted a deep bidirectional transformer architecture (BERT) to perform our multi-label classification task [33]. We utilized a pre-trained BERT model for the Russian language DeepPavlov [34]. We tuned the last layer of the model, which consisted of 12 sigmoid neurons. As a baseline, we used a long-short term memory (LSTM) ANN, which consisted of embedding as the first layer and one LSTM layer (100 cells), dropout (*p* = .2) and a subsequent multi-label dense layer with sigmoid as the activation function.

### Model evaluation

We trained the BERT and LSTM models using a stratified k-fold validation scheme where *k* = 5. Because the classes were imbalanced, we utilized an up-sampling strategy; that is, underrepresented classes were up-sampled in the training dataset. The testing set distribution was not modified. Table 1 displays the evaluation results. Precision and recall were calculated for both micro- and macro-averaged aggregations [35]. As shown in Table 1, precision and F1 scores were reported for thresholds equal to 0.5. We utilized a computer with a Tesla T4 GPU to train the models. Table 1 shows that BERT outperformed the LSTM model by a large margin. We therefore chose the BERT model and trained it on 95% of the data; in this case, it returned a micro-averaged accuracy of 0.94 and an area under the ROC curve (AUC) score of 0.991.

**Table 1:**
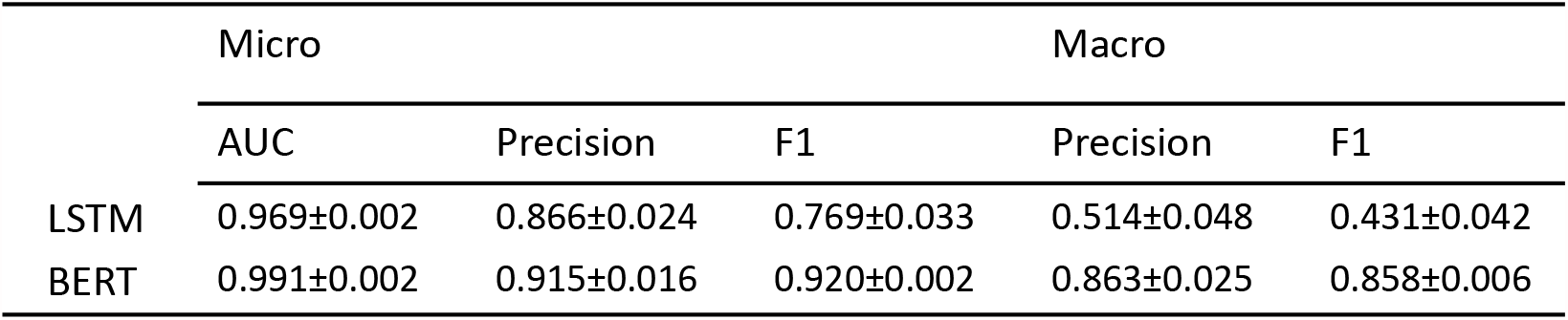
BERT and LSTM models evaluation results.

Regarding gender, age, and dose number (if available), we used counts of corresponding abbreviations and regular expression matching because the administrators of the group had provided detailed instructions for the reporting of this information.

## Results

Reactogenicity assessment based on opt-in civic surveillance was performed to obtain results of clinical importance (similar to endpoints in trials).

### Temporal Dynamics

The volume peak of the self-reports corresponded to the time at which vaccinations were speeded up (Figure 1). Moreover, after three months of vaccinations (the end of February 2021), the popularity of self-reporting started to decrease despite the increasing vaccination roll-out. However, the Pearson correlation coefficient between doses administered until February 28 was very high (*r* = .7, P<.001) and the subsequent count of administered doses increased, while reports on AEs decreased (Figure 1).

**Figure 1:**
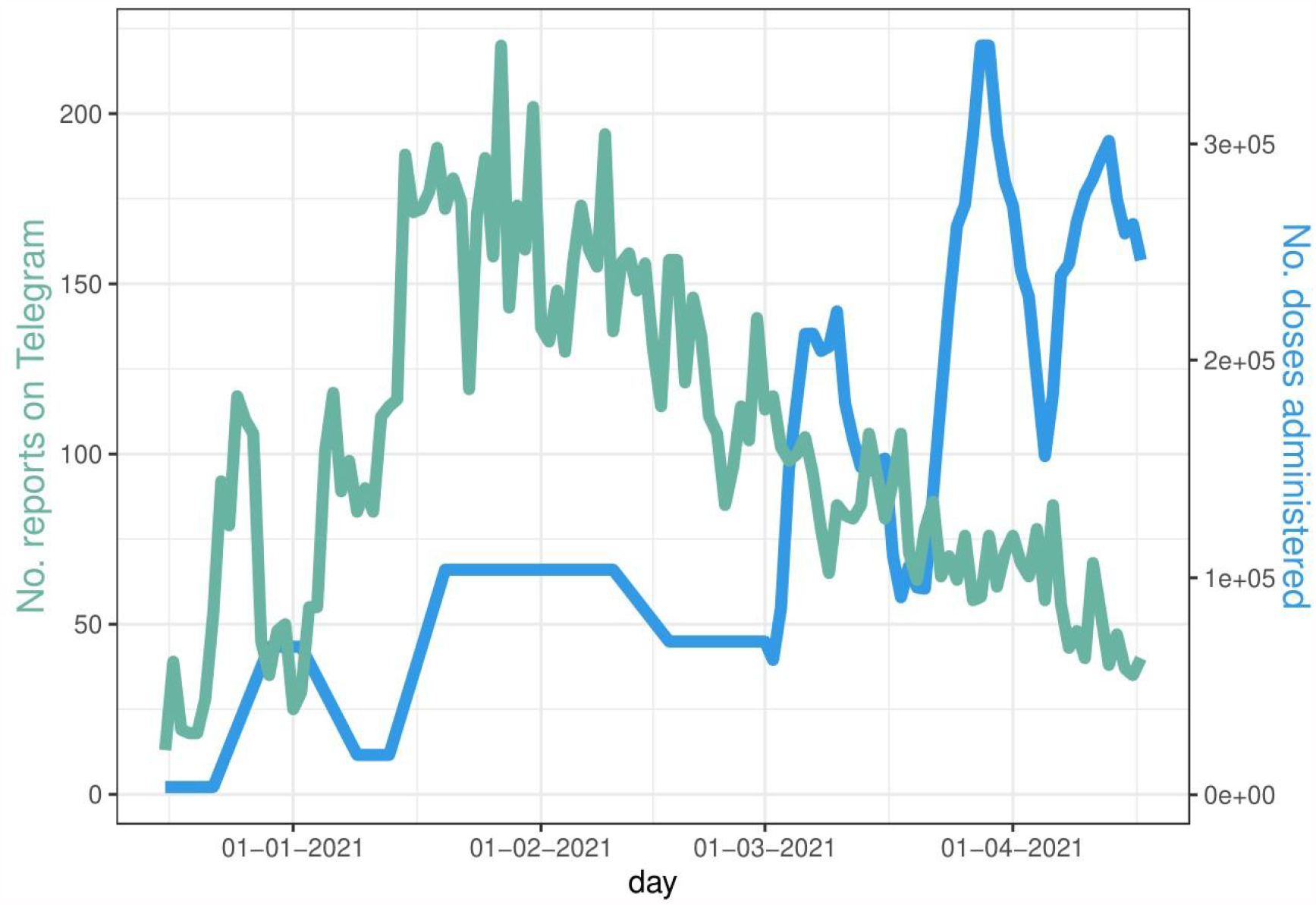
Daily counts of reports of AE and doses administered in Russia (data according to Our World in Data [36])

### Revealed AE Frequencies (BERT classes)

Our analysis revealed that fever and generalized pain were the most commonly reported AEs (Table 2, Figure 2). Injection site irritations (local reactions) were in the order of magnitude less likely to be reported than fever and pain (systemic reaction). Gastric symptoms (especially diarrhea, with a frequency of 0.6% per report) were less likely to be reported than the average prevalence among the general population (1–5% for diarrhea).

**Table 2:** Frequencies of mild adverse events extracted from the Telegram group

|  | fever | pain | insomnia | fatigue | nausea/<br>vomiting | headache | erythema/<br>redness | pruritus | swelling | lymph nodes<br>enlargement | diarrhea | chills |
| --- | --- | --- | --- | --- | --- | --- | --- | --- | --- | --- | --- | --- |
| Frequency | 47.4% | 46.6% | 5.2% | 33.5% | 3.0% | 24.8% | 2.7% | 1.7% | 1.8% | 1.6% | 0.6% | 23% |

**Figure 2:**
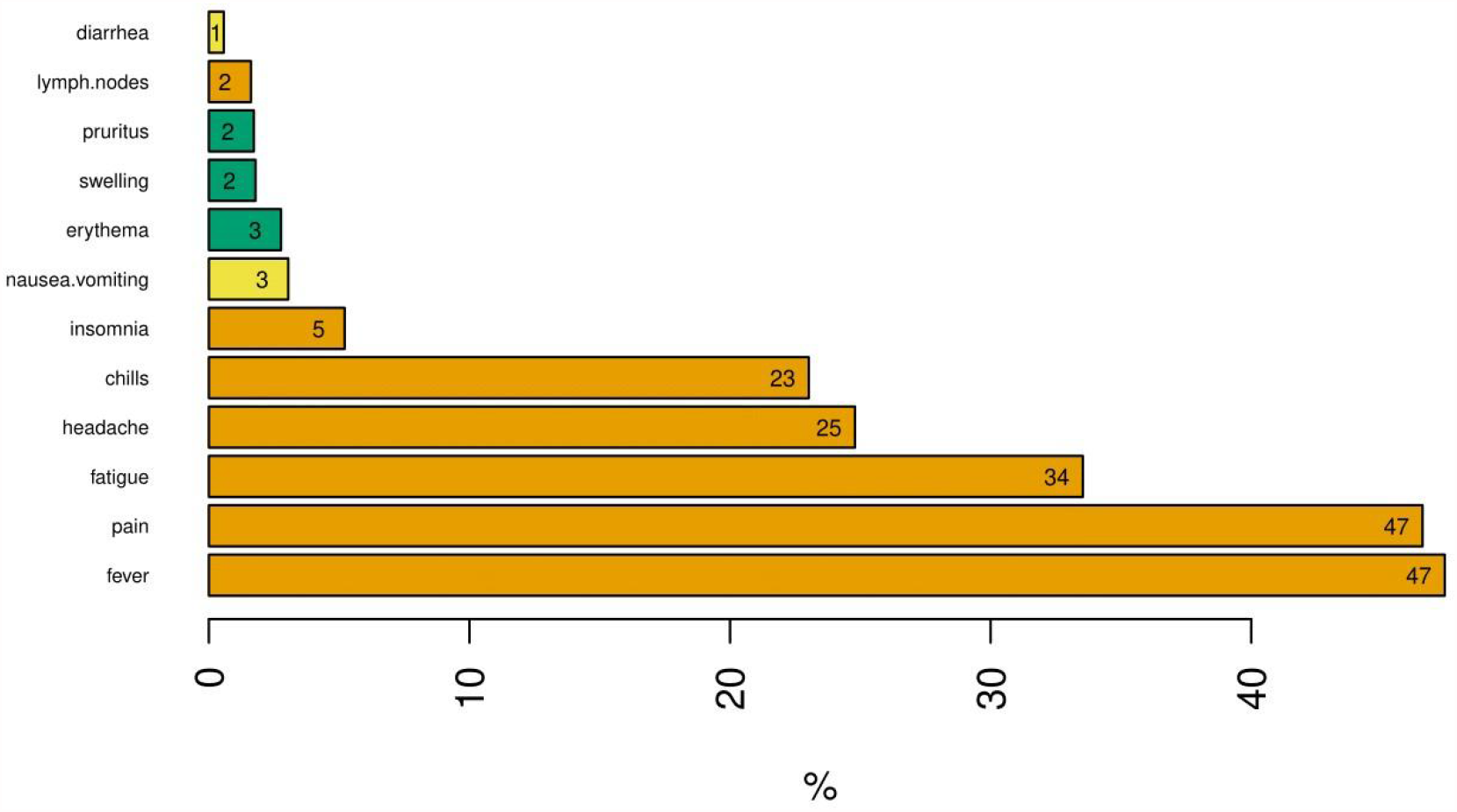
Frequency (%) of given AE classified to be reported by users. Symptoms: orange – systemic; green – local; yellow – gastric

### Variations across Age, Gender, and Dose

Gender was reported by 3,992 females and 2,762 males. On average, females reported 2.5 AEs (*σ* = 1.79), and males reported 2.1 AEs (*σ* = 1.64). Females reported statistically significantly more AEs (*P* = 2 · 10^−16^) according to the results of a Mann-Whitney U test (Table 3).

**Table 3:** Comparisons of mean number of AEs in various groups by gender and by dose and Mann-Whitney U test results. OR stands for Odds Ratio.

| gender | mean<br>no. AEs | OR | P-value | dose | mean<br>no. AEs | OR | P-value |
| --- | --- | --- | --- | --- | --- | --- | --- |
| male | 2.1 | 1.20 | $2 \cdot 10^{-16}$ | first | 2.2 | 1.13 | $1.7 \cdot 10^{-5}$ |
| female | 2.5 |  |  | second | 1.9 |  |  |

Age was provided by 6,754 users. A linear regression was performed for those who reported being at least 18 years old (minimal age of Russian registration [1]). We found a clear and significant linear relation, showing that with every year of life, the users reported *β* = .0457 + */* − .0014 AE less. Mild AEs among the elderly are known to be less frequently observed for most anti-COVID vaccines [29–31,37].

AEs in response to other anti-COVID-19 vaccines have been found to depend on whether the vaccination was the first or the second dose (if applicable). For instance, AEs in response to mRNA vaccines have tended to be stronger with the second dose [30,31,37]. In contrast, AEs involving vector vaccines have tended to be milder with the second dose [38]. Regarding the Sputnik V vaccine, this difference might be because a different vector is used in each dose, which might lead to different reactions. Among the self-reports, 4,174 described AEs after the first dose, 1,251 after the second dose, and 3,049 described AEs after both doses. It is also possible that the users did not receive the second dose because of contraindications or just lost interest in reporting.

Here, we considered only reports that discussed the first and second doses separately. On average, there were 2.2 (*σ* = 1.80) AEs for the first dose and 1.9 (*σ* = 1.69) AEs for the second dose. According to the results of the Mann-Whitney U test, there were statistically significantly more AEs after the first dose (*P* = 1.7 · 10^−5^) (Table 3).

### Co-occurrence of AEs

To quantify the co-occurrence of symptoms, we calculated Spearman’s rank correlation coefficients between each pair of classified symptoms. We observed systemic, local, and gastric clusters (Figure 4). We also provided a network representation in which vertex size represents symptom prevalence and edge width represents co-occurrence as measured by the correlation coefficient. Only edges with a correlation coefficient above 0.09 are shown (Figure 4). An unsupervised weighted Louvain algorithm for community detection was used for this purpose, and the vertices were colored the same if they belonged to the same community, which revealed a meaningful structure in which orange denoted systemic, green denoted local, and yellow denoted gastric communities of symptoms.

**Figure 3:**
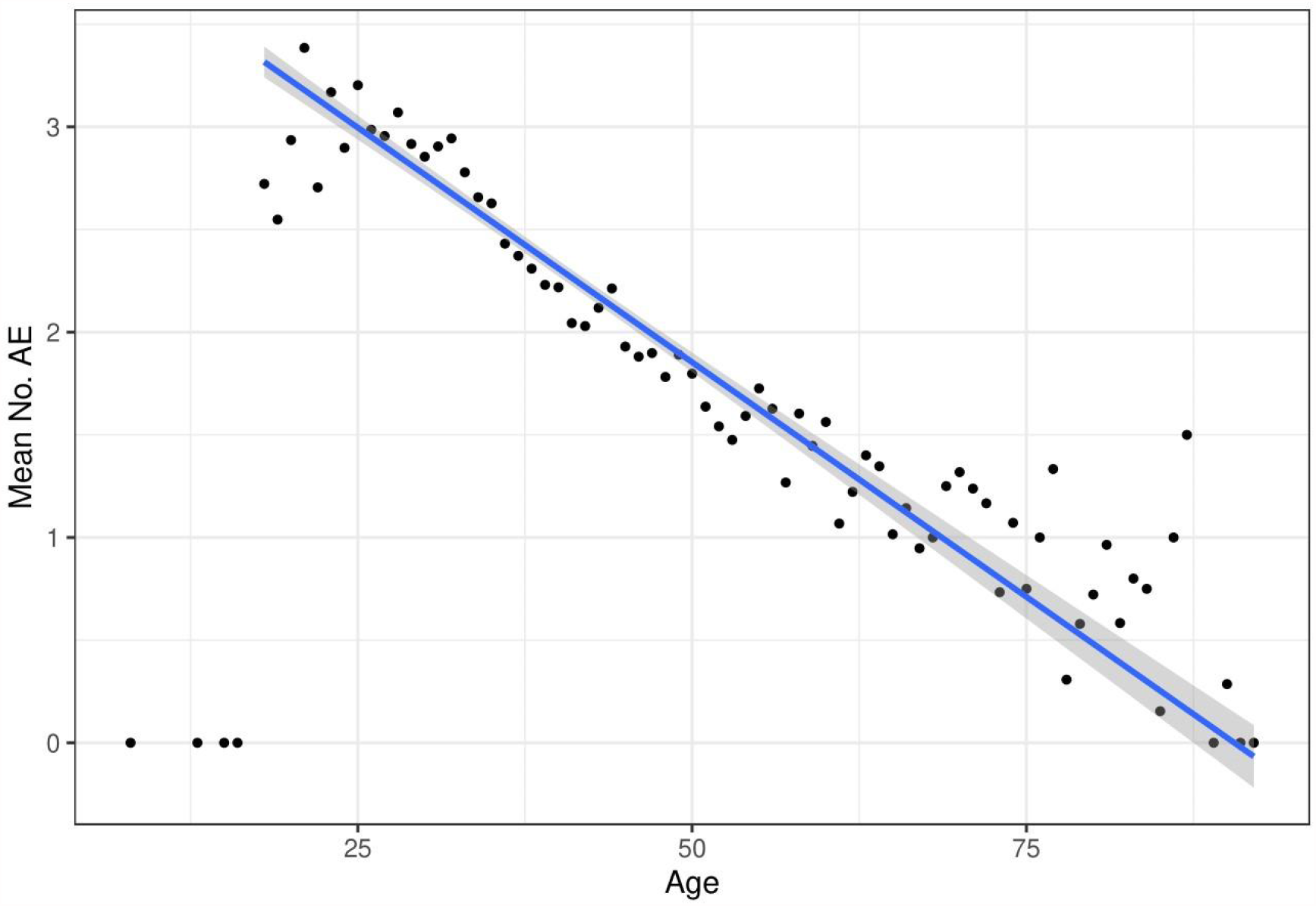
Scatter plot of age against number of AEs reported. Dots indicate the mean number of AEs for a given age. The blue line indicates a linear regression trend with confidence intervals. Some artifacts were observable (e.g., reported age less than 18 years).

**Figure 4:**
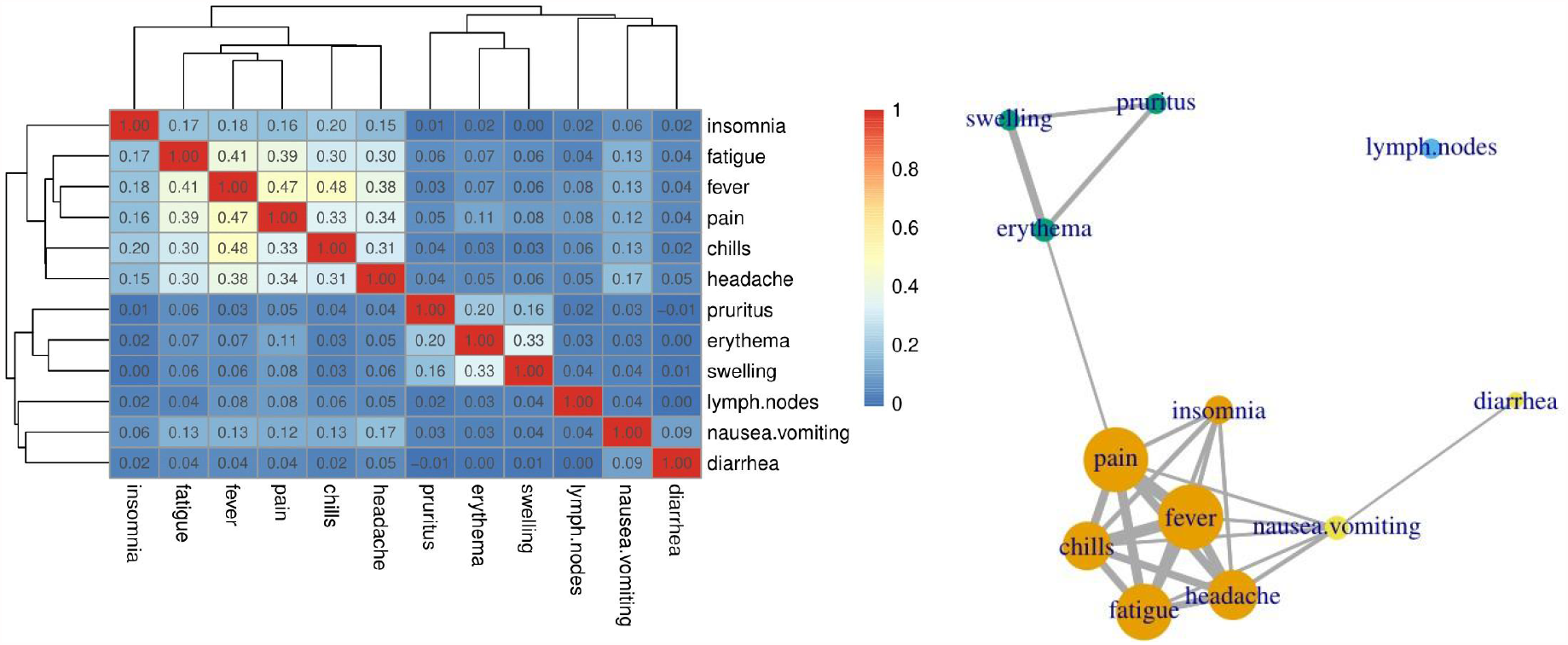
AEs co-occurrence: (left) Hierarchical correlation matrix for AE symptoms and (right) community structure of network of AE symptoms.

### Telegram Versus Other Trials or Registries of Sputnik V

We compared our results with two available datasets of AEs in response to the Sputnik V vaccine. The first one was collected in Moscow. The second one was collected in Argentine.

#### Moscow clinical trial

Mild AEs in 1,029 patients older than 60 years in phase III of the clinical trial [40] were compared with 690 self-reports by Telegram users older than 60 years. Because there were inconsistencies in various definitions of AEs, a simplified classification was provided, and only headache and diarrhea comprised similar symptoms (at least *sensu lato*).

We performed the following calculations to compare both datasets. To obtain *fever* according to our definition, we summed over *pyrexia, fever sensation*, and *elevated body temperature* from the clinical trial. Similarly, to obtain *pain*, we summed over *myalgia, arthralgia, and local reaction*. To obtain *fatigue*, we summed over *asthenia* and *malaise*. To obtain *nausea*, we summed over *nausea* and *dyspepsia*. For *erythema*, we chose *contact dermatitis*.

In all systemic reactions, Telegram users reported AEs significantly more often than measured in the clinical trial (Table 4). In contrast, diarrhea was less likely to be reported than measured in the clinical trial.

**Table 4:** Frequency (%) of given category in Telegram and clinical trial data sets of Sputnik AE [40] with Pearson correlation coefficients comparing both samples. Fisher’s test was applied for the comparison of each symptom between samples (with given *p*-values). The reference to the odds ratio is for a clinical trial in Moscow.

| Correlation<br>(*r* = .69, *P* = .08) | pain | headache | fatigue | fever | nausea | erythema | diarrhea |
| --- | --- | --- | --- | --- | --- | --- | --- |
| Clinical trial | 6.70 | 2.90 | 3.00 | 3.20 | 1.20 | 3.80 | 0.80 |
| Telegram | 25.65 | 12.90 | 20.43 | 23.62 | 1.30 | 2.17 | 0.43 |
| Odds ratio | 3.82 | 4.42 | 6.78 | 7.59 | 1.12 | 0.57 | 0.56 |
| *p*-Value | $4.7 \cdot 10^{-21}$ | $1.9 \cdot 10^{-13}$ | $1.1 \cdot 10^{-26}$ | $3.5 \cdot 10^{-32}$ | 0.83 | 0.09 | 0.54 |

#### Argentinian post-registration AE registry

Another available dataset on AEs in response to Sputnik V was compiled from the Argentinian registry of AE monitoring. This registry contains 23,804 events of all kinds (96.5% = mild AEs) according to 2,541,362 doses administered. To compare, we chose 7,797 Telegram posts that reported at least one AE, and we adjusted new disjoint subsets of symptoms according to the Argentinian methodology [41].

We categorized gastric as the frequency of logical function nausea OR diarrhea. We categorized site irritation as the frequency of logical function pruritus OR erythema OR swelling. We categorized fever_pain as the frequency of logical function fever AND (pain OR headache). We categorized fatigue_pain as the frequency of logical function fatigue AND (pain OR headache). We categorized only_fever as the frequency of logical function fever AND ∼(pain OR headache OR fatigue); ∼ denotes logical negation.

The comparison showed that the statistics, despite the significant differences shown in Table 5, were similar in magnitude and highly correlated (r = 0.94). The comparison of the Telegram reports (a selected sample with at least one AE constructed by multi-label classification) with the Argentinian registry (multi-class classification [41]) was conducted by the mapping described above. The results of the comparison must be interpreted with caution.

**Table 5:** Frequency (%) given category in Telegram and Argentinian safety monitor [41] for Sputnik V AEs. Fisher’s test was applied for each comparison of each symptom between samples (with given *p*-values). The reference to the odds ratio is to the Argentinian registry. Pearson correlation coefficients comparing the series of symptoms in both samples are provided.

| Correlation ($r = .94$, $P=.02$) | fever_pain | fatigue_pain | gastric | site irritation | only_fever |
| --- | --- | --- | --- | --- | --- |
| Argentinian Registry | 33.20 | 38.10 | 5.98 | 9.30 | 8.3 |
| Telegram | 54.70 | 39.67 | 5.14 | 7.31 | 9.53 |
| Odds ratio | 1.66 | 1.05 | 0.88 | 0.80 | 1.12 |
| p-Value | $1.2 \cdot 10^{-91}$ | 0.06 | 0.03 | $2.5 \cdot 10^{-6}$ | 0.02 |

### Comparison with Other Vaccines

Regarding vaccines registered by the EMA and FDA, lists of the frequency of the most common adverse events are accessible; however, they vary across regulatory bodies. Thus, we chose a subset of symptoms for frequencies that were reasonably comparable (pain, headache, fatigue, fever, chills, and nausea). We built a distance (Euclidean) matrix of AEs based on clinical trial registries (EMA [38, 42–44], FDA [29–31]) and from the Telegram group. From the FDA dataset, for two-dose vaccines, the dose with higher reactogenicity was selected. In clinical trials, pain is usually considered pain at the injection site. Fever was the sum of pyrexia and fever in the EMA database. EMA used the injection site tenderness/irritation category. However, regarding redness/erythema, the FDA classified swelling and pruritus separately. Thus, erythema was not included. Sputnik V is a vector vaccine, as are AstraZeneca and Johnson & Johnson. The results showed that Telegram Sputnik V AEs were clustered with other vector vaccines, which was possibly due to similar safety profiles (Figure 5).

**Figure 5:**
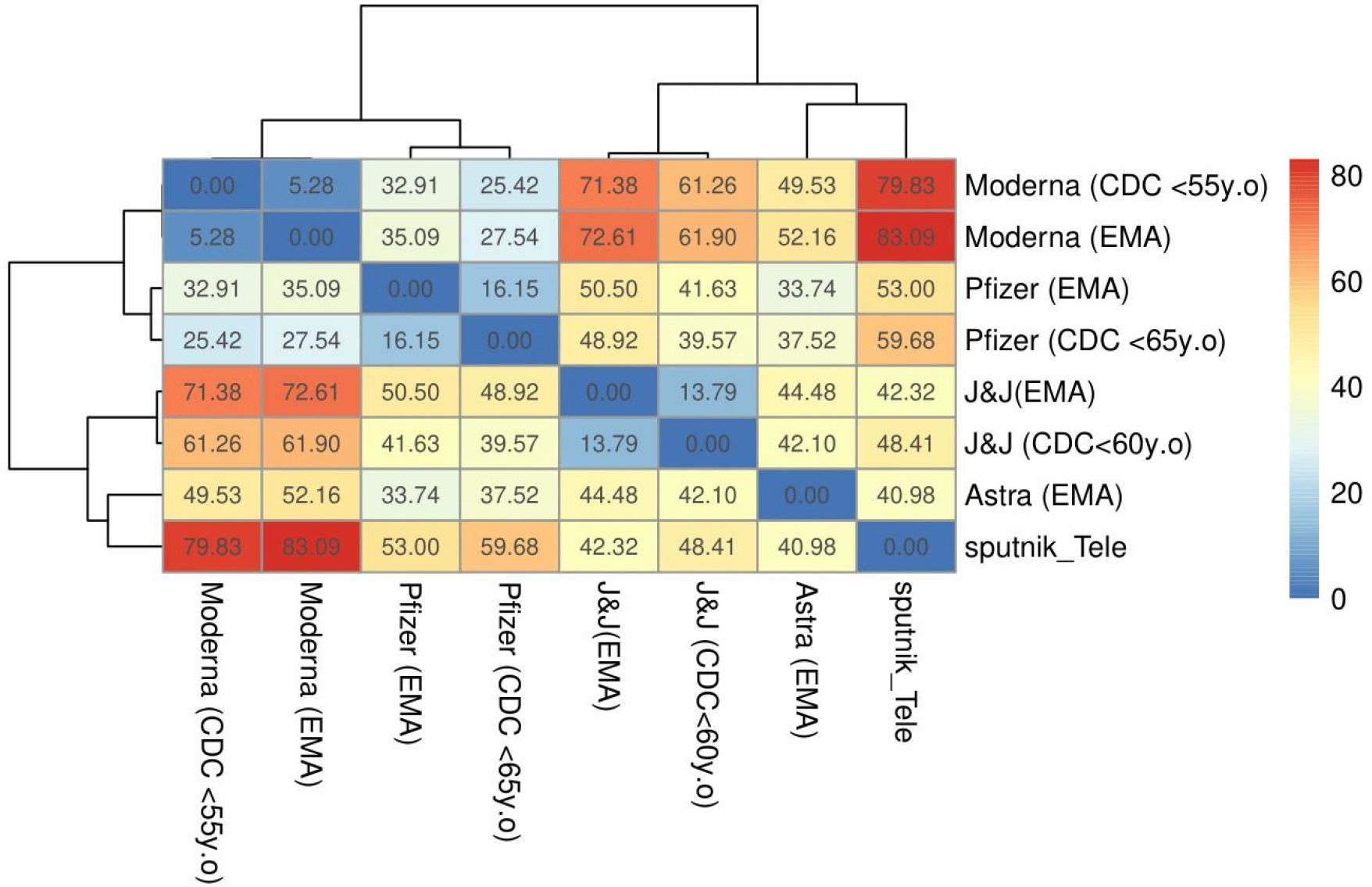
Hierarchical matrix of various vaccines and reporting systems (Euclidean distance) of vaccinations investigated in the present study. Astra stands for AstraZeneca, J&J stands for Johnson & Johnson.

It is important to note that the Telegram users also submitted reports without any AEs at all. Thus, our surveillance system included a sentinel property of samples in contrast to VAERS (North America), ARR (European Union), and the Argentinian registry [41], which gather reports only if there is any AE to be reported.

## Discussion

Mild, non-severe AEs have usually been ignored by medical communities because they are common to all vaccines. Antivax movements have emphasized severe AEs, which have been widely discussed in social media [45] in the wider context of vaccine safety [46,47]. In the discourse on COVID-19 vaccines, the main issues were that they were developed quickly, and they could compromise safety. Those issues included the fear that vaccines would alter human DNA, cause allergic reactions to vaccine ingredients, result in sudden deaths due to frailty syndrome, or cause infertility [48,49]. Wide anti-COVID immunization programs promulgated a discourse in which risk (e.g., the discomfort of common, but mild AEs as well as rare, but serious AEs) and benefits (e.g., efficacy in protecting from the disease) were described as “tradeoffs” of being vaccinated. Mild AEs have become an important issue for many people; moreover, they have the economic component of the potential need for sick leave. This discourse led to the formation of a public Telegram group, where users were asked to report AEs.

In the present study, we demonstrated that in the first phase of the vaccination roll-out the AE reports were correlated (correlation r = .7) with vaccination volume (Figure 1). However, the Telegram users tended to lose interest after a few months. It is possible that because of the prioritization of vaccine delivery, which began with public and military servants, scientists, teachers, and medical staff, these “early adopters” were more likely to post on social media and be actively involved in reporting AEs. Subsequently, users in the general population were vaccinated, and they were less involved in reporting on the Telegram platform (Figure 1).

The results of the present study showed that the number of reported AEs decreased linearly according to age (*β* = .05 AE per year; Figure 3). This result was dependent on biology, which was confirmed in previous clinical trials [30,31,42] and post-marketing observations [37] of other anti-COVID-19 vaccines. Telegram users older than 60 years reported significantly more systemic AEs compared with their peers in clinical trials, who tested negative for having or having recovered from COVID-19 [40] (Table 4). On one hand, it is possible that people previously infected with COVID-19 were more likely to report AEs after receiving other vaccines [37]. On the other hand, self-reporting bias could be an important factor in explaining the difference between the “Moscow’s” clinical trial and the Telegram reports.

The safety profile of the Sputnik V vaccine includes mild AEs, which is more similar to vector vaccines than to mRNA anti-COVID vaccines, which was quantified by the Euclidean distance between AE frequencies (Figure 5). Sputnik V safety profile also showed a high fever to fatigue ratio (Table 6) and a stronger reaction to the first dose than to the second one (Table 3).

**Table 6:** Frequency (%) of AEs in response to Sputnik V (Telegram) and other vaccines (EMA and CDC/FDA)

|  | pain | headache | fatigue | fever | chills | nausea |
| --- | --- | --- | --- | --- | --- | --- |
| AstraZeneca (EMA) | 54.20 | 52.60 | 53.10 | 41.50 | 31.90 | 21.8 |
| Johnson & Johnson (EMA) | 48.60 | 38.90 | 38.20 | 14.00 | 5.00 | 14.2 |
| Johnson & Johnson (CDC < 60 yrs.) | 59.80 | 44.40 | 43.80 | 12.80 | 5.00 | 15.5 |
| Pfizer (EMA) | 80.00 | 50.00 | 60.00 | 30.00 | 30.00 | 5.00 |
| Pfizer (CDC < 65 yrs.) | 77.80 | 51.70 | 59.40 | 15.80 | 35.10 | 10.00 |
| Sputnik (Telegram) | 46.57 | 24.80 | 33.54 | 47.43 | 23.02 | 3.00 |
| Moderna (CDC < 55 yrs.) | 90.10 | 62.80 | 67.60 | 17.40 | 48.30 | 21.3 |
| Moderna (EMA) | 92.00 | 64.70 | 70.00 | 15.50 | 45.40 | 23.00 |

Women reported more AEs than men did (1.2-fold, *P <* 0.001 Mann-Whitney U test). This phenomenon is well recorded in other anti-COVID-19 vaccine registries [37,50], which could indicate sex-dependent vaccine reactivity. However, this result needs to be understood with caution. The CDC has warned that gender bias in reporting could be more important than possible biological mechanisms [37]. The likelihood of disclosing personal information (even anonymously) is known to vary, such as according to gender [51], social class [52] and so forth. A potential reason is that women are more likely to be interested in health, write about health on the Internet, and disclose their information [51].

On Telegram, self-reports are most likely to underestimate gastric symptoms (e.g., diarrhea 0.6%). These symptoms could be a taboo effect [53], such as a response to public speaking anxiety. Alternatively, it could be easily ignored because of its high prevalence, or it could be eliminated using an over-the-counter medicine such as loperamide [54,55]. Insomnia was detected so often that it suggests an epidemiological link with the vaccine, which needs further investigation. Local AEs, such as injection site irritation, have rarely been reported. Underlying conditions of erythema/redness, which is usually one of the most common AEs in responses to all injected substances, including vaccines, are probably overlooked due to low subjective discomfort and lack of physical investigation by a doctor. The findings showed that their actual prevalence was probably underreported.

Our study has several limitations. First, we analyzed participatory/community-based surveillance among Russian Telegram users. Therefore, the results may be specific to the Russian population in a given stage of the pandemic and therefore should not be extrapolated to other contexts. Second, Telegram users may overlook less troublesome side effects, and the social context could influence decisions on taking part in discussions and being selective in reporting AEs [51,52]. For example, local or gastric AEs could be underreported. Third, the classifications developed in this study should not be strictly applied in other contexts. For example, pain at the injection site and pain in other parts of the body were not distinguished. Finally, because our infodemiology study focused on community research initiatives (independent and nonprofit projects, with already known strengths and weaknesses from the history of medicine [56]), our observations cannot replace real-world studies [57–59].

## Conclusion

The symptoms reported by social media users only partially reflect their prevalence in the real world [60]. Therefore, the frequencies of symptoms should not be interpreted without considering the contexts and proportions of other symptoms (i.e., fever to fatigue ratio), the phase of the epidemic, and the vaccination roll-out (i.e., the number of doses administered daily and the population that is vaccinated as willingness to report AEs satisfy typical product life-cycle temporal characteristics [60]).

According to clinical trials [40] and official registries [41], only partial information could be retrieved on Sputnik V. Previously, multiple researchers have raised concerns about the safety of the Sputnik V vaccine [6,57,58]. Our study aimed to increase transparency regarding the safety and efficacy of Sputnik V [59] like the US-based early warning objective of actively surveilling COVID-19 vaccine safety through v-safe [61].

In the present study, we showed that community-based surveillance via social media (so-called civil “clinical trials”) can provide meaningful information that could be useful, and this phenomenon should be carefully investigated. The frequencies of AEs extracted from Telegram samples in which at least one AE was reported were in line with the official Argentinian data on passive surveillance (r = .94 correlation). Because the authenticity and credibility [14] of the reports were not assessed in our study, it is likely that incorrect information was included in the data. Nonetheless, social media monitoring is promising for gathering information on common mild AEs of not only vaccines but also other pharmaceutical products [62].

In the future, it would be of interest to explore the extraction of AEs from a dataset using named entity recognition methods that consider noisy data [11] and to apply nested NER methods [63].

## Data Availability

Raw data available upon request

## Acknowledgements

The authors acknowledge the creators and users of the Telegram group “Sputnik_results”.

## Footnotes

1 https://t.me/Sputnik_results

2 https://www.adrreports.eu

3 https://vaers.hhs.gov

4 https://yellowcard.mhra.gov.uk

## References

1. Государственный реестр лекарственных средств, Гам-КОВИД-Вак Комбинированная векторная вакцина для профилактики коронавирусной инфекции. https://grls.rosminzdrav.ru/Grls_View_v2.aspx?routingGuid=6c1f7501-7067-45b3-a56d-95e25db89e97&t 2020. Accessed: 2021-03-21.

2. Logunov DY, Dolzhikova IV, Zubkova OV, et al. Safety and immunogenicity of an rAd26 and rAd5 vector-based heterologous prime-boost COVID-19 vaccine in two formulations: two open, non-randomised phase 1/2 studies from Russia. Lancet. 2020;396(10255):887–897.

3. Logunov DY, Dolzhikova IV, Shcheblyakov DV, et al. Safety and efficacy of an rAd26 and rAd5 vector-based heterologous prime-boost COVID-19 vaccine: an interim analysis of a randomised controlled phase 3 trial in Russia. Lancet. 2021;397(10275):671–681.

4. ourworldindata. Most Approve of National Response to COVID-19 in 14 Advanced Economies. https://ourworldindata.org/coronavirus 2021. Accessed: 2021-03-21.

5. FT. Russia seeks to make Sputnik V in Italy as overseas demand surges. https://www.ft.com/content/905ee381-ef16-4fa1-ac38-a8b2bb2df16f 2021. Accessed: 2021-03-21.

6. Baraniuk C. Covid-19: What do we know about Sputnik V and other Russian vaccines?. BMJ. 2021;372.

7. Karafillakis E, Martin S, Simas C, et al. Methods for Social Media Monitoring Related to Vaccination: Systematic Scoping Review. JMIR public health and surveillance. 2021;7(2):e17149.

8. Samaras L, Garc’ sıa-Barriocanal E, Sicilia MA. Syndromic surveillance using web data: a systematic review. Innovation in Health Informatics. 2020:39–77.

9. Zheluk A, Gillespie JA, Quinn C. Searching for Truth: Internet Search Patterns as a Method of Investigating Online Responses to a Russian Illicit Drug Policy Debate. J Med Internet Res. 2012;14(6):e165.

10. Sboev A, Sboeva S, Gryaznov A, Evteeva A, Rybka R, Silin M. A neural network algorithm for extracting pharmacological information from russian-language internet reviews on drugs. in Journal of Physics: Conference Series;1686:012037IOP Publishing 2020.

11. Dai X, Karimi S, Paris C. Medication and adverse event extraction from noisy text. in Proceedings of the Australasian Language Technology Association Workshop 2017:79–87 2017.

12. Salehan M, Kim DJ. Predicting the performance of online consumer reviews: A sentiment mining approach to big data analytics. Decision Support Systems. 2016;81:30–40.

13. Gattepaille LM, Hedfors Vidlin S, Bergvall T, Pierce CE, Ellenius J. Prospective evaluation of adverse event recognition systems in Twitter: Results from the Web-RADR project. Drug safety. 2020;43:797–808.

14. Hoang T, Liu J, Pratt N, et al. Authenticity and credibility aware detection of adverse drug events from social media. International journal of medical informatics. 2018;120:157–171.

15. Adrover C, Bodnar T, Huang Z, Telenti A, Salath’ se M. Identifying adverse effects of HIV drug treatment and associated sentiments using Twitter. JMIR public health and surveillance. 2015;1(2):e7.

16. Zhou Z, Hultgren KE. Complementing the US Food and Drug Administration Adverse Event Reporting System With Adverse Drug Reaction Reporting From Social Media: Comparative Analysis. JMIR Public Health and Surveillance. 2020;6(3):e19266.

17. Li Y, Jimeno Yepes A, Xiao C. Combining Social Media and FDA Adverse Event Reporting System to Detect Adverse Drug Reactions. Drug safety. 2020;43:893–903.

18. Patel R, Belousov M, Jani M, et al. Frequent discussion of insomnia and weight gain with glucocorticoid therapy: an analysis of Twitter posts. NPJ digital medicine. 2018;1(1):1–7.

19. De Castro NML, Samart’ sın-Ucha M, Mart’ın-Vila A, Alvarez-Payero M, Pin∼eiro-Corrales G,’ Pego-Reigosa JM. Content analysis of Twitter in relation to biological treatments for chronic inflammatory arthropathies: an exploratory study. European Journal of Hospital Pharmacy. 2019;26(3):124–128.

20. Smith K, Golder S, Sarker A, Loke Y, O’Connor K, Gonzalez-Hernandez G. Methods to compare adverse events in Twitter to FAERS, drug information databases, and systematic reviews: proof of concept with adalimumab. Drug safety. 2018;41(12):1397–1410.

21. Pierce CE, Bouri K, Pamer C, et al. Evaluation of Facebook and Twitter monitoring to detect safety signals for medical products: an analysis of recent FDA safety alerts. Drug safety. 2017;40(4):317–331.

22. Huesch MD. Commercial online social network data and statin side-effect surveillance: a pilot observational study of aggregate mentions on facebook. Drug safety. 2017;40(12):1199–1204.

23. Semenov A, Mantzaris AV, Nikolaev A, et al. Exploring Social Media Network Landscape of Post-Soviet Space. IEEE Access. 2019;7:411–426.

24. Statista. Leading social media platforms in Russia as of 3rd quarter of 2020, by penetration rate. https://www.statista.com/statistics/867549/top-active-social-media-platforms-in-russia/ 2021 Accessed: 2021-03-05.

25. Wang J, Zhao L, Ye Y, Zhang Y. Adverse event detection by integrating twitter data and VAERS. Journal of biomedical semantics. 2018;9(1):1–10.

26. Jarynowski A. Sputnik V Adverse Events risk calculator, https://infodemia-koronawirusa.shinyapps.io/sputnik/ 2021.. Accessed: 2021-06-05.

27. Telegram. Telegram Terms of Service. https://telegram.org/tos 2021. Accessed: 2021-03-05.

28. Github T. Telethon Github. https://github.com/LonamiWebs/Telethon 2021. Accessed: 2021-03-05.

29. CDC. Local Reactions, Systemic Reactions, Adverse Events, and Serious Adverse Events: Janssen COVID-19 Vaccine. https://www.cdc.gov/vaccines/covid-19/info-by-product/janssen/reactogenicity.html 2021. Accessed: 2021-05-01.

30. CDC. Local Reactions, Systemic Reactions, Adverse Events, and Serious Adverse Events: Moderna COVID-19 Vaccine. https://www.cdc.gov/vaccines/covid-19/info-by-product/moderna/reactogenicity.html 2020. Accessed: 2021-03-21.

31. CDC. Local Reactions, Systemic Reactions, Adverse Events, and Serious Adverse Events: Pfizer-BioNTech COVID-19 Vaccine. https://www.cdc.gov/vaccines/covid-19/info-by-product/pfizer/reactogenicity.html 2020. Accessed: 2021-03-21.

32. LabelStudio. LabelStudio. https://labelstud.io/ 2021. Accessed: 2021-03-05.

33. Devlin J, Chang MW, Lee K, Toutanova K. BERT: Pre-training of Deep Bidirectional Transformers for Language Understanding. 2019.

34. MIPT. DeepPavlov GitHub. https://github.com/deepmipt/DeepPavlov 2021. Accessed: 2021-04-05.

35. Sokolova M, Lapalme G. A systematic analysis of performance measures for classification tasks. Information Processing Management. 2009;45(4):427–437.

36. Mathieu E, Ritchie H, Ortiz-Ospina E, et al. A global database of COVID-19 vaccinations. Nature Human Behaviour. 2021.

37. Menni C, Klaser K, May A, et al. Vaccine side-effects and SARS-CoV-2 infection after vaccination in users of the COVID Symptom Study app in the UK: a prospective observational study. The Lancet Infectious Diseases. 2021.

38. EMA. COVID-19 Vaccine AstraZeneca -SUMMARY OF PRODUCT CHARACTERISTICS. https://ec.europa.eu/health/documents/community-register/2021/20210129150842/anx_150842_en.pdf 2021. Accessed: 2021-03-21.

39. Blondel VD, Guillaume JL, Lambiotte R, Lefebvre E. Fast unfolding of communities in large networks. Journal of statistical mechanics: theory and experiment. 2008;2008(10):P10008.

40. Logunov DY, Dolzhikova IV, Tukhvatullin AI, Shcheblyakov DV. Safety and efficacy of the Russian COVID-19 vaccine: more information needed–Authors’ reply. The Lancet. 2020;396(10256):e54–e55.

41. Ministerio Salud. Campaña Nacional de Vacunación contra la COVID-19. 10° Informe de vigilancia de seguridad en vacunas 2 de abril code 2021.. https://bancos.salud.gob.ar/recurso/10deg-informe-de-seguridad-en-vacunas 2021. Accessed: 2021-05-01.

42. EMA. COVID-19 Vaccine Janssen-SUMMARY OF PRODUCT CHARACTERISTICS. https://www.ema.europa.eu/en/documents/product-information/covid-19-vaccine-janssen-epar-product-information_pl.pdf 2020. Accessed: 2021-05-01.

43. EMA. COVID-19 mRNA Vaccine Moderna-SUMMARY OF PRODUCT CHARACTERISTICS. https://ec.europa.eu/health/documents/community-register/2021/20210106150575/anx_150575_en.pdf 2020. Accessed: 2021-03-21.

44. EMA. COVID-19 mRNA Vaccine Comirnaty-SUMMARY OF PRODUCT CHARACTERISTICS. https://www.ema.europa.eu/en/documents/product-information/comirnaty-epar-product-information_en.pdf 2020. Accessed: 2021-03-21.

45. Klimiuk K, Czoska A, Biernacka K, Balwicki L. Vaccine misinformation on social media–topic based content and sentiment analysis of Polish vaccine-deniers’ comments on Facebook. Human Vaccines & Immunotherapeutics. 2021:1–10.

46. Kata A. Anti-vaccine activists, Web 2.0, and the postmodern paradigm–An overview of tactics and tropes used online by the anti-vaccination movement. Vaccine. 2012;30(25):3778–3789.

47. Cianciara D, Szmigiel A. Posting on „Nie szczepimy (“We don’t vaccinate”) Internet forum. Przegląd epidemiologiczny. 2019;73(1):105–115.

48. Griffith J, Marani H, Monkman H. COVID-19 Vaccine Hesitancy in Canada: Content Analysis of Tweets Using the Theoretical Domains Framework. J Med Internet Res. 2021;23(4):e26874.

49. Thelwall M, Kousha K, Thelwall S. Covid-19 vaccine hesitancy on English-language Twitter. Profesional de la informacion (EPI). 2021;30(2).

50. Gee J. First month of COVID-19 vaccine safety monitoring—United States, December 14, 2020– January 13, 2021. MMWR. Morbidity and mortality weekly report. 2021;70.

51. Elnegaard S, Andersen RS, Pedersen AF, et al. Self-reported symptoms and healthcare seeking in the general population-exploring “The Symptom Iceberg”. BMC public health. 2015;15(1):1–11.

52. Sæbø Ø, Federici T, Braccini AM. Combining social media affordances for organising collective action. Information Systems Journal. 2020.

53. Kowalski P. O tym, co nieuniknione: ekskrementy i defekacja. in Colloquia Anthropologica et Communicativa;4:19–41 2011.

54. Kamiński M, Borger M, Prymas P, et al. Analysis of Answers to Queries among Anonymous Users with Gastroenterological Problems on an Internet Forum. International journal of environmental research and public health. 2020;17(3):1042.

55. Panayiotou G, Karekla M, Georgiou D, Constantinou E, Paraskeva-Siamata M. Psychophysiological and self-reported reactivity associated with social anxiety and public speaking fear symptoms: Effects of fear versus distress. Psychiatry research. 2017;255:278–286.

56. Epstein S. Impure science: AIDS, activism, and the politics of knowledge. Univ of California Press; 1996.

57. Tulleken C. Covid-19: Sputnik vaccine rockets, thanks to Lancet boost. bmj. 2021;373.

58. Vlassov V. Covid-19: What do we know about Sputnik V and other Russian vaccines?. bmj opinion. 2021.

59. Logunov DY, Dolzhikova IV, Shcheblyakov DV. Data discrepancies and substandard reporting of interim data of Sputnik V phase 3 trial–Authors’ reply. Lancet (London, England). 2021.

60. Rogers EM. Diffusion of innovations. Simon and Schuster 2010.

61. Shimabukuro TT, Kim SY, Myers TR, et al. Preliminary Findings of mRNA Covid-19 Vaccine Safety in Pregnant Persons. New England Journal of Medicine. 2021.

62. Freifeld CC, Brownstein JS, Menone CM, et al. Digital drug safety surveillance: monitoring pharmaceutical products in twitter. Drug safety. 2014;37(5):343–350.

63. Strakova J, Straka M, Hajic J. Neural Architectures for Nested NER through Linearization. in Proceedings of the 57th Annual Meeting of the Association for Computational Linguistics (Florence, Italy):5326–5331 Association for Computational Linguistics 2019.

